# Simulation of epidemic models with generally distributed sojourn times

**DOI:** 10.1101/2021.01.15.21249880

**Authors:** Carlos Hernandez-Suarez, Osval Montesinos-Lopez, Ramon Solano-Barajas

**Affiliations:** Facultad de Ciencias, Universidad de Colima, Bernal Diaz del Castillo 340, Colima, Colima, 28040, MEXICO; Facultad de Telematica, Universidad de Colima, Av. Universidad 333, Colima, Colima, 28040, MEXICO; Facultad de Ingenieria Civil, Universidad de Colima, Carr. Colima-Coquimatlan, Km 9, Coquimatlan, Colima, 28400, MEXICO

**Keywords:** Stochastic, Epidemic models, Markov chains, Simulations, Sojourn time

## Abstract

Epidemic models are used to analyze the progression or outcome of an epidemic under different control policies like vaccinations, quarantines, lockdowns, use of face-masks, pharmaceutical interventions, etc. When these models accurately represent real-life situations, they may become an important tool in the decision-making process. Among these models, compartmental models are very popular and assume individuals move along a series of compartments that describe their current health status. Nevertheless, these models are mostly Markovian, that is, the time in each compartment follows an exponential distribution. Here, we introduce a novel approach to simulate general stochastic epidemic models that accepts any distribution for the sojourn times.

## 1. Introduction

A mathematical model is a real-life sketch that allows for experimentation and testing. Engineers have used models for long time, where temperature, friction, durability and forces play a role in decision making. Similarly, chemists have used mathematical models to analyze chemical reactions with the purpose of optimization, but, despite the long-standing tradition of using mathematical models, their use in biology or medicine is much more recent. Sometimes scientists in these disciplines deal with not well understood phenomena and thus, the set of assumptions is usually larger and require the conjunction of different disciplines. From the classical Ross Malaria model (Ross, 1915) that allowed to conclude that keeping the mosquito population below a threshold would eliminate Malaria, to Blower’s findings (Blower and McLean, 1994) that a vaccine with a low level of protective efficacy may do more harm than good by providing a sense of false security to the vaccinated, models may reveal a hidden ‘cobra effect’, that is, something that has not accounted for.

Mathematical models have been a valuable tool to analyze the demographics of biological populations and have become a valuable tool to biologists to follow the health of populations in the field (Caswell, 2001). As Cohen states: “*Mathematics is biology’s next microscope, only better; biology is mathematics’ next physics, only better*” (Cohen, 2004).

Epidemics are not only driven by etiological agents but also by the behavior of the population. For this later, mathematical models in epidemiology resemble more a field of economics than of medicine. Examples of such behaviour may involve the population’s reaction to vaccination, abortion, use of face-masks, medication, blood or plasma transfusions, condom use, etc. The fact that a model considers such behaviors does not guarantee that these have been correctly included in the model, which most of the times requires a deep knowledge of the phenomena and interdisciplinary work. For instance, Needle/Srynge Policies (NSOs) that attempt to reduce HIV transmission by providing needles for free, must face the fact that sharing needles is sometimes part of the experience (Kaplan and Heimer, 1994), and that peer pressure is a determinant factor in acquiring smoking habits (Evans et al., 1978).

The most popular epidemic models consist of a series of stages that represent the different real-life health status or conditions. Individuals move along the compartments according some specified transition rules. The analysis of these models can be deterministic or stochastic. There are many differences between these two types of models, but here we illustrate the two that are the most relevant. In Figure 1 we can see a stage model with three compartments, *X, Y* and *Z*. The deterministic model consists of a continuous *drainage* of individuals from all compartments and thus, if at time *t* = 0 there is one individual in compartment *X*, after some finite time *t*, the individual will be spread among the three compartments. In contrast, in a stochastic model, the individual can be in only one of them. The most important difference is the source of uncertainty: in deterministic models, based on differential equations, there is no uncertainty, whereas in stochastic models, the outcome may vary under the same parameter set. Figure 2 shows 5 stochastic simulations of an SEIR epidemic model shown together with the solution of the deterministic model. Clearly, the distribution of the stochastic simulations is shifted to the right, since small epidemics vanish quickly and are unnoticeable. This will result in differences in the average time to extinction, the cost of the epidemics (the integral under the curve of infected) and other functionals of time.

**Figure 1:**
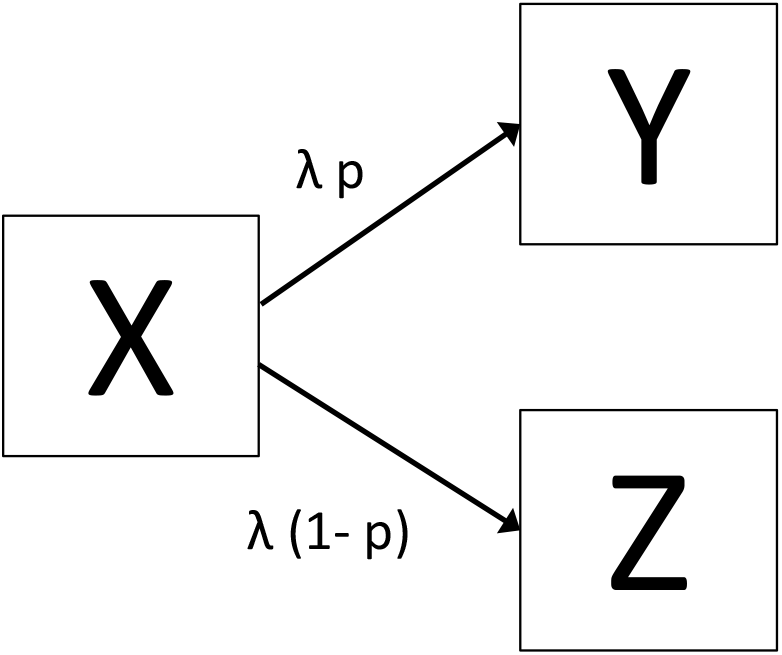
The main differences between deterministic and stochastic models can be shown in this Figure. Suppose that at time *t* = 0 there is only one individual in compartment *X*. In a deterministic model this individual is *drained* continuously at a rate *λ* and a fraction *p* of this goes to compartment *Y*, thus, after some finite time *t*, this individual is spread among the three compartments, whereas in a stochastic model the individual is in only one of them.

**Figure 2:**
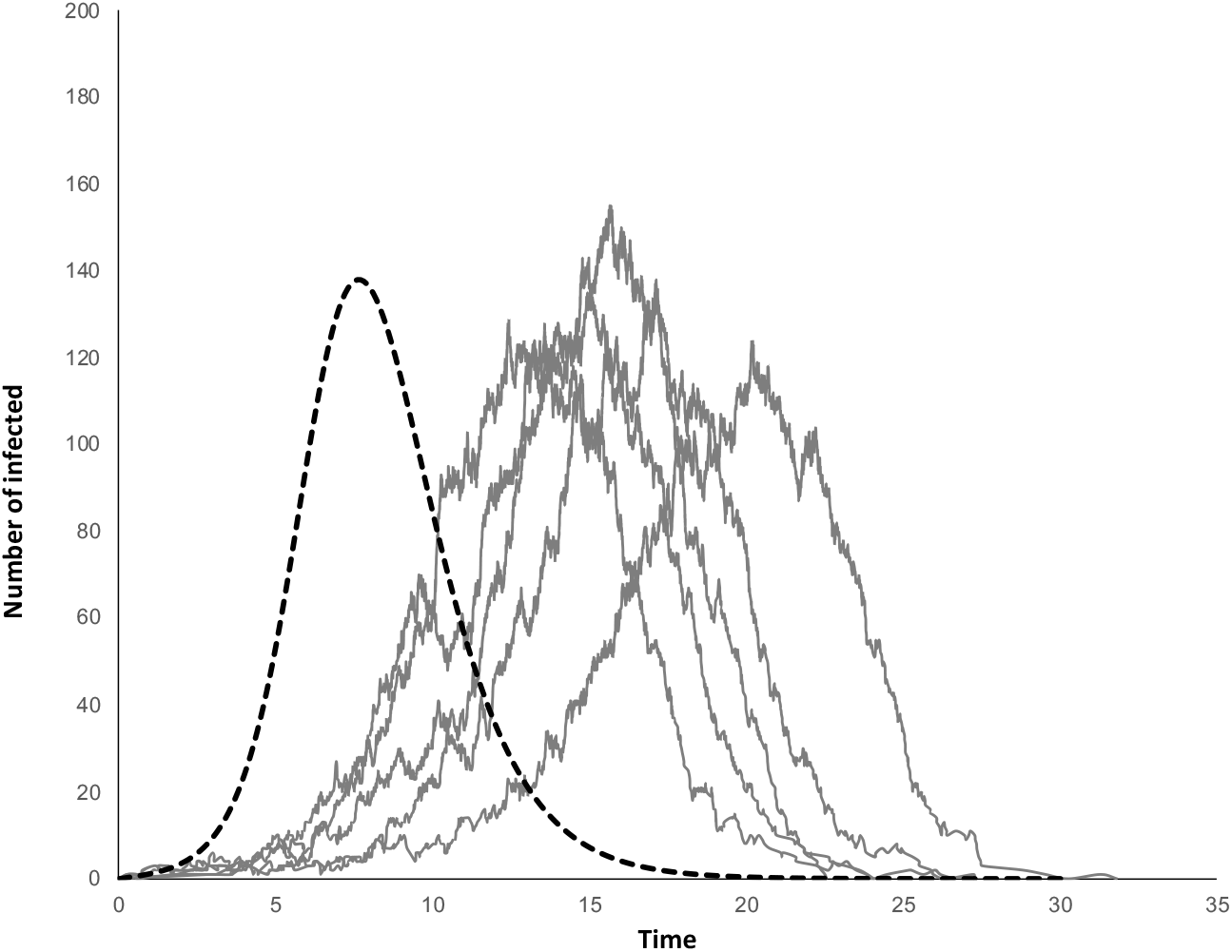
Stochastic simulations of as SEIR model. Some simulations vanished quickly and are unnoticeable. The plot shows the number of infectious (I) over time, with *N* = 1000 and *R*_0_ = 2. Dashed line is the solution of the deterministic model.

## 2. Simulations

Before the advent of computers one could only rely on analytic results which limited the complexity of the models. Computer simulations allow researchers to obtain the response to *‘What would happen if*…*’* in complex situations. In deterministic models, the term *simulation* refers to the numerical solution of a set of differential equations. In stochastic models, it implies the use of pseudo-random numbers to decide when an individual leaves a compartment as well as the origin and destination compartments. Stochastic models allow researchers to obtain information on the variability of the possible outcomes: while in a deterministic model the number of infected at time *t* is constant, in stochastic models, this number follows a statistical distribution whose moments may be infered by performing a large number of simulations.

In Markovian epidemic models, sojourn times in each compartment are exponential random variables. Simulating these models is easy because the *memoryless property* allows to describe the whole system at any time *t* with a vector containing the number of individuals in each compartment. But the mean and standard deviation of an exponential distribution are equal, which is a very stringent condition for real-life situations. If sojourn times are not exponential we would need to keep track no only of the number of individuals in any given compartment, but also on the actual time each individual has been there, which affects the decision on who moves next and where to. Keeping this record is computationally intensive.

We first provide a review of what is being called as *time to step* simulation, used in simulating epidemic models with exponential sojourn times and how it is commonly dealt when the sojourn time is not exponential.

### Review of “time to step” simulation

As mentioned before, whereas in a deterministic model there is a continuous ‘leaking’ between boxes, in the Markovian version of the model, only one transition between two compartments takes place at any time, which is a random event and the time when this transition takes place is another (independent) continuous random variable. The following results from exponential distributions are used in *time to step* simulations:

If *X* and *Y* are two exponential distributions with respective parameters *α* and *β*, then:

i. *P* (*X < Y*) = *α/*(*α* + *β*)
ii. The min(*X, Y*) follows an exponential distribution with parameters *α* + *β*
iii. The probability that *X < Y* is independent of the distribution of min(*X, Y*).
iv. If an event occurs at a time which follows an exponential distribution with parameter *α* but when the event occurs, there is a probability *p* that event will be of type *A*, then the time to the occurrence of an event of type *A* follows an exponential distribution with parameter *αp*.

Property (iii) implies that the time to the next event is independent of the event that will take place. This is an important property because we can independently simulate the time to the next event and the event that will take place. Property (iv) is called *thinning* the Poisson processes.

Assume that there are *K* compartments and that transitions at time *t* from compartment *i* to compartment *j* occur a a rate *δ*_*ij*_(*t*). For simplification, we will write this rate as *δ*_*ij*_. A stochastic simulation of these models involves:

a. Calculating the rates of all transitions *δ*_*ij*_.
b. Calculating the total transition rate *R* = ∑_*ij*_ *δ*_*ij*_.
c. Calculating the transition probabilities *p*_*ij*_ for every possible transition: as *p*_*ij*_ = *δ*_*ij*_*/R*.
d. Simulating when the next transition will take place using an exponential distribution with parameter *R*.
e. Selecting the transition that will take place.
f. Updating the system, that is, moving one individual from compartment *i* to compartment *j* according the chosen transition in step (e) and adding the simulated time obtained in step (d).
g. Re-starting from step (a) and continuing until *R* = 0 or a predefined stopping time *t*_max_.

Due to property (iii) steps (e) and (d) can be exchanged. For step (d), since the time to the next transition is exponentialy distributed, we can simulate it by using *T* = *−log*(*u*)*/R*, where *u* is a uniform random number (0, 1). For step (e), we can select a single sample from a multinomial distribution with parameters *p*_*ij*_, for *i* = 1, 2, …, *K*; *j* = 1, 2, …, *K* where the outcome is the index of the event that will take place.

Tthis kind of simulation is known as *time to event* simulation because we modify the contents in each compartment until the next event takes place. Thus, in a SEIR model the number of times the loop (a)-(g) above has to be executed is about 3 times the number of infected, since each one of them must move eventually through the stages *E, I* and *R*. For *R*_0_ large, the number of infected may be close to *N*, the population size.

### Review of fitting non-exponential sojourn times to the compartments

As mentioned before, stochastic Markovian models assume that the duration of time in each compartment is exponential because these distributions have the *memoryless property* in which regardless of the time an individual has been in a compartment, the probability that the individual will leave it in the next *s* units of time is 1 *− e*^*−λs*^, for some *λ*. Consequently, it is not necessary to track the current time of an individual in a compartment to decide if it is time to leave it or not. We only need to know how many individuals there are in each compartment and the respective exit rates to decide what happens next. Exponential times implies a constant hazard rate, which is not precisely a characteristic of most stages in most diseases. For instance, an individual that just had surgery may present complications from surgery, and the possibility of a complication in the next unit of time reduces with time, thus, this is an example of a decreasing hazard rate. On the other hand, an individual that has been infected may eventually die or recover eventually and the possibility that one of these events will occur in the next unit of time increases with time, thus, it has a positive hazard rate.

An attempt to approximate a distribution with some mean *µ* and some variance *σ*^2^ can be done using Erlang distributions (Anderson and Watson, 1980; Lloyd, 2001; Wearing et al., 2005; Champredon et al., 2018). This approach is equivalent to divide a compartment into several sub-compartments, each with an exponential duration, so that the sojourn time through all of them has the desired mean and variance. An exact match is sometimes impossible and we need to get as close as we can to the desired moments. There are two problems inherent to using this technique: the first one is that, in a model with *K* stages (excluding the susceptible state), there are roughly *K −*1 transitions for each individual, and if each of those *K* stages is divided in *n* substages, the number of transitions increases by *n*, resulting in more computing time. The second problem is that the approximation of the first two moments of a single stage alone may require a very large number of substages. For instance, if we want to match a distribution with mean 20 days and standard deviatio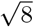, we would need an Erlang distribution with parameters 50 and 5*/*2, that is, 50 compartments in a row, each with an exponential sojourn time with parameter 5*/*2. At some point, the number of compartments may become prohibitive, and some relaxations would be made, and the distribution is only approximated. If this is repeated for a model with several compartments, things can become very complicated.

## 3. A new method to simulate stochastic compartmental models

In most epidemic models, an individual starts in a compartment *S* for the susceptible stage and then starts a passage through some or all of the remaining stages. Once an individual leaves the *S* compartment, the passage through the other stages is independent of what other individuals are doing. Assume that there are *N* individuals and *K* stages excluding the susceptible, thus, we can generate a matrix **T** of size *N × K* with entries *t*_*i,j*_ for the duration of individual *i* in compartment *j*. Since most popular computer scientific programs can handle *vectorization* to generate random variables, the generation of each column of **T** is fast.

The only problem that remains is deciding when an individual becomes infected, if this occurs. Let **y** a vector of size *N ×* 1 where *y*_*i*_ is the time when individual *i* leaves the *S* compartment. The vector **y** and the matrix **T** contain all information of one simulation. We start with one infected at time *t* = 0, that is, *y*_1_ = 0, whose transition through the rest of the stages is contained in the first row of **T**. Vector **y** is not easy to get, and is, by far, the most time-consuming step in the process.

Sellke (1983) developed an alternative construction of a stochastic SIR epidemic that we will use to derive vector **y**. Sellke’s goal was to provide a simple proof of Daniel’s (1967) result regarding the final number of removed, that is, the size of an epidemic. Sellke’s derivation relaxed the small number of initially infected, but still needed the assumption of an exponential distributed infectious period. Here, Ball (1986) version of Sellke’s construction is used because it does not require the assumption of exponential distribution for the duration of the infectious state.

If the population is of size *N* and there is an initial infected at time *t*_0_, according to Sellke (1983) and Ball (1986), the *exposure* required to achieve infection follows a Poisson process with parameter *λ/N*. Therefore, to generate a vector **L**^*′*^ = [*L*_1_, *L*_2_, *L*_3_, …, *L*_*N−*1_] with the ordered required exposures, we must first generate a sorted vector **u** = [*u*_1_, *u*_2_, *u*_3_, …, *u*_*N−*1_] where *u*_*i*_ follow a Uniform distribution in [0, 1] such that *u*_*i*_ *> u*_*i*+1_ and then:

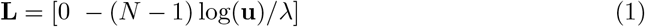

The *i*-th individual will leave the *S* compartment after reaching a level of exposure equal to *L*_*i*_, but the exposure required by individual *i* + 1 is equal to the duration of infection of the *i* previously infected, consequently, if this sum is smaller than *L*_*i*+1_, the *i* + 1-th individual will not be infected and the epidemic stops with *i* infected. Figure 3 illustrates the idea behind this construction. The epidemic terminates with *n* infected when:

**Figure 3:**
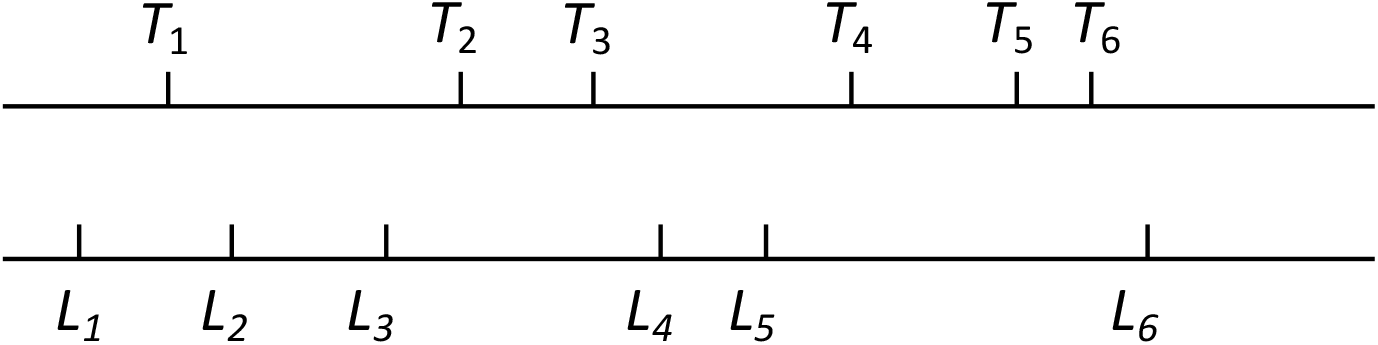
Sellke’s construction of an epidemic starts with a single infected whose infectious period is *T*_1_. Intervals *T*_*i*_ on the top line correspond to the duration of the infectious period of the individuals in the order they were infected, whereas the bottom line *L*_*i*_, represent the amount of exposure required to achieve infection by the *i*-th individual. The sixth infection requires an exposure greater than 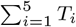, therefore, the epidemics stops with 5 infected. This construction of the epidemic is useful to calculate the final epidemic size but neither *L*_*i*_ nor *T*_*i*_ represent the actual times of infection or removal and these must be calculated from the *T*_*i*_’s and *L*_*i*_’s. *T*_*i*_’s are not required to be exponential distributions.

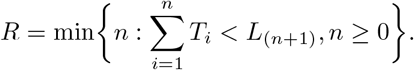

Although there are many ways to generate vector **y** containing the times when individuals leave the susceptible state, we chose a general approach that can be used with more complicated models, for instance, models with several infectious stages with differential degree of infectiousness. This approach is based on the concept of *individual exposure* that we will describe.

### 3.1. Individual exposure, f_j_(y_j_, t)

The *individual exposure* of individual *j, f*_*j*_(*y*_*j*_, *t*), that is, the force of infection produced by this individual, is a piecewise function defined as follows: let **W** be the cumulative sum over rows of matrix **T**. Let 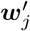 be the *j*-th row of **W** appending a 0 at the beginning, that is:

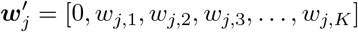

The individual exposure *f*_*j*_(*y*_*j*_, *t*) is:

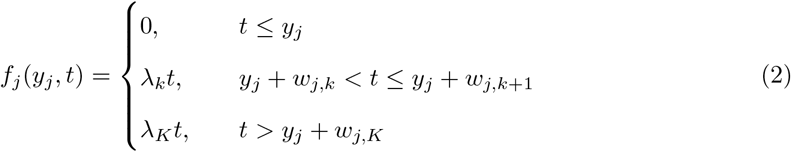

where *λ*_*k*_ is the infection rate of individuals in stage *k*. As we can see, *y*_*j*_ in *f*_*j*_ (*y*_*j*_, *t*) is the time when the *j*-th individual leaves the *S* compartment. Two examples of *individual exposures* are depicted in Figures 4a and 4b.

**Figure 4:**
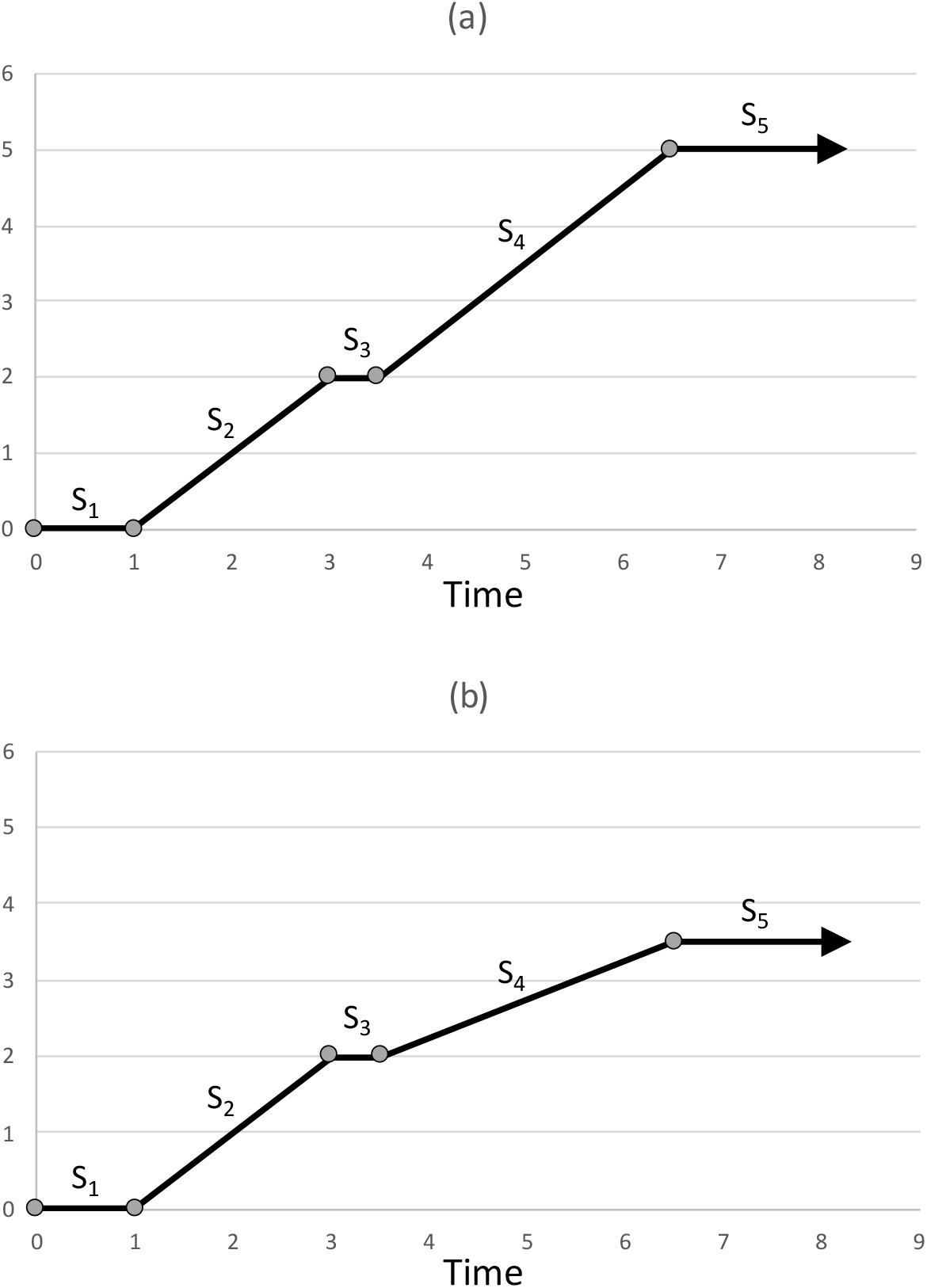
The *individual exposure f*_*j*_ (0, *t*). In both cases the sojourn time in each of the five stages *S*_1_, *S*_2_, *S*_3_, *S*_4_, *S*_5_ are: [1, 2, 0.5, 3, 1.8]. In (a) the contact rates are [0, 1, 0, 1, 0] whereas in (b) these are [0, 1, 0, 0.5, 0], that is, stage *S*_4_ is half infectious in the second case, resulting in a smaller individual exposure. The arrows at the end of both lines indicate the exposure remains constant starting stage 5.

Let ***λ***^*′*^ = [*λ*_1_, *λ*_2_, …, *λ*_*K*_] a vector with the infectious contact rates of each stage. Define ***E*** = ***W λ*** and similarly let 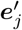 be the *j*-th row of **E** appending a 0 at the beginning, that is:

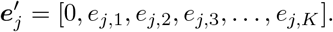

Observe that *f*_*j*_ (*y*_*j*_, *t*) is completely defined by the set 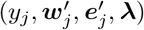, where 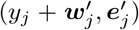 marks the boundaries of each segment, with slopes contained in ***λ***.

### 3.2. The accumulated exposure, F_j_ (t)

After *j* infections, the infected individuals have provided some exposure to infection which is a non-decreasing piecewise function *F*_*j*_ (*t*) as the one depicted in Figure 5. *F*_*j*_ (*t*) is also a piecewise set of linear functions with inflection points when some individual started or stopped as infectious. Given *j* infections, the infection of the next individual starts at 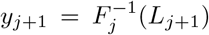 and therefore has an individual exposure *f*_*j*_ (*y*_*j*+1_, *t*). To update the exposure to *F*_*j*+1_(*t*), we need to perform:

**Figure 5:**
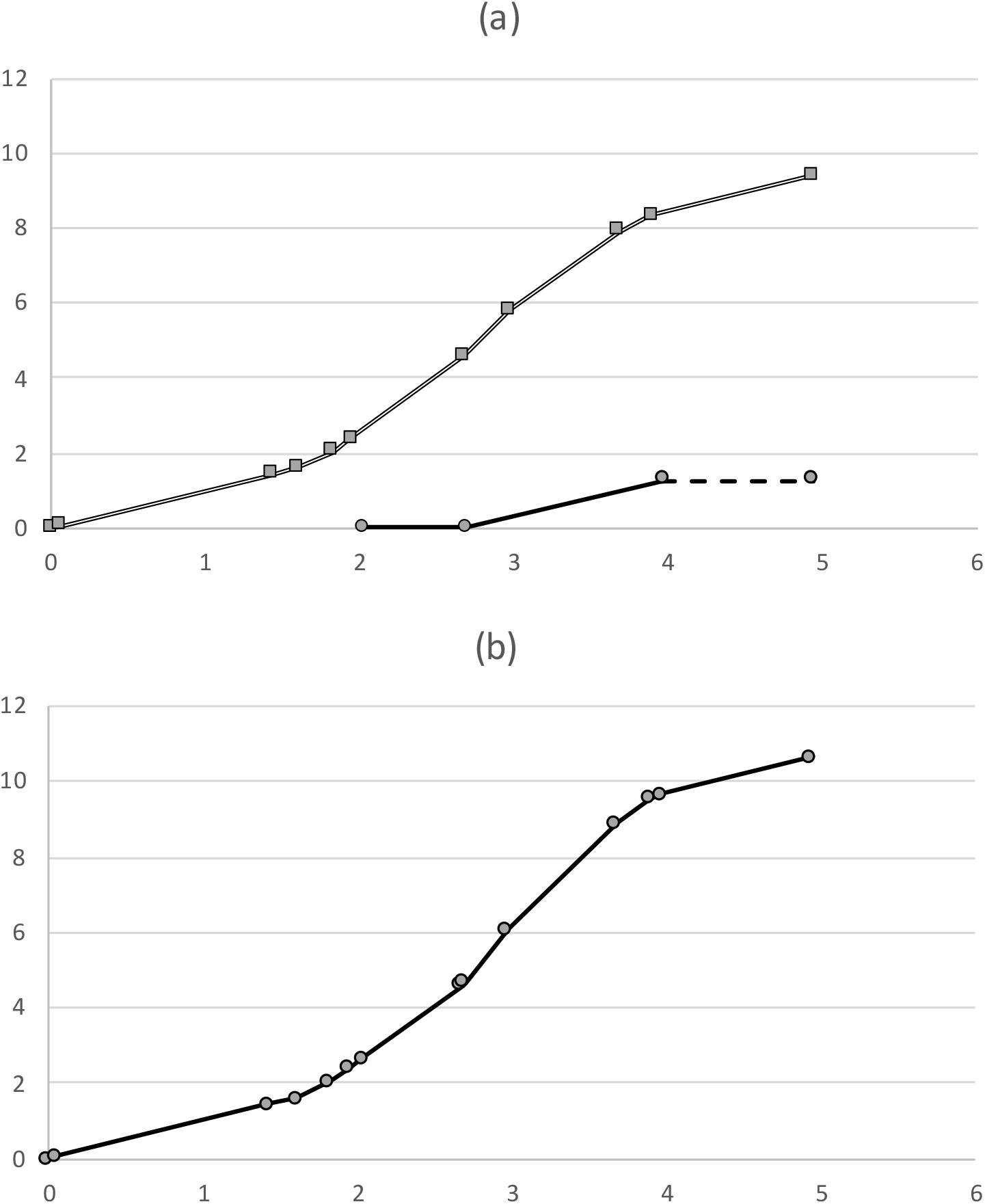
The top line in Figure (a), is *F*_*j*_ (*t*), the accumulated exposure achieved by the previous *j* infectious individuals. The (*j* + 1)-th infection starts at 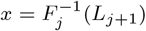 where *L*_*j*+1_ is the exposure required by the *j* + 1 individual. Once the starting point is derived, we update the accumulated exposure by adding *F*_j+1_(*t*) and the new individual exposure. In this example the line in the bottom of Figure (a) is a new infection of an individual that required an exposure of 2.66, thus, it starts at *t* = 2.02 This individual will remain latent for 0.67 units of time and then it will start its infectious period that will last 1.26 units. The dashed line is the accumulated exposure provided by this individual. Figure (b) shows both lines in Figure (a) after being added, producing the updated exposure.

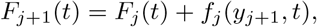

with *F*_0_(*t*) = 0. Figure 5 illustrates the idea to calculate when the next infection will take place.

### 3.3. Dealing with several infectious stages with different contact rates

When there is a single infectious stage with contact rate *λ*, the exposure required by each individual to be infected can be simulated using (1). When there are *r* infectious stages with differential degree of infectiousness we have a vector of contact rates ***λ***^*′*^ = [*λ*_1_, *λ*_2_, …, *λ*_*K*_]. Choose some positive and finite *λ*^*∗*^ and let ***λ***^*′*^ = *λ*^*∗*^[*r*_1_, *r*_2_, …, *r*_*K*_] where *r*_*i*_ = *λ*_*i*_*/λ*^*∗*^. The exposure required for the individuals to become infected is obtained with **L** = *−*(*N −* 1) log(**u**)*/λ*^*∗*^. Clearly, the exposures required contained in **L** depends on the chosen *λ*^*∗*^. Nevertheless, the time to infection is not affected by the chosen *λ*^*∗*^ because the shape of *individual exposures* change (Figure 5). This is the essence of the Time-scale Transformation Algorithm to simulate random events from a non-homogeneous Poisson process. We provide a simple proof in the Appendix.

### 3.4. A suggested pseudocode to obtain the times of infection y

The pseudocode to obtain the infection times is actually very simple:

i. Let ***λ***^*′*^ = [*λ*_1_, *λ*_2_, *λ*_3_, …, *λ*_*K*_] be the infectious contact rates of the different stages.
ii. Choose some *λ* positive and finite so that ***λ***^*′*^ = *λ*[*k*_1_, *k*_2_, *k*_3_, …, *k*_*r*_], a vector with the infectious contact rates of the different stages.
iii. Generate required *N −* 1 exposure times as **L** = *−*(*N −* 1) log(**u**)*/λ*, where ***u*** is a vector of size *N −* 1 of uniform (0, 1) random numbers. Append a 0 to the beginning of ***L***.
iv. Set *j* = 1. Start with *f*_*j*_(0, *t*) and let *F*_*j*_(*t*) = *f*_*j*_(0, *t*)
v. Set *j* = *j* + 1. Let 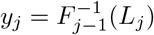.
vi. Generate *f*_*j*_(*y*_*j*_, *t*) as in (2) and obtain *F*_*j*_(*t*) = *F*_*j−*1_(*t*) + *f*_*j*_(*y*_*j*_, *t*)
vii. Return to (v), stop with *j* infected if there is no *y*_*j*+1_ that satisfies 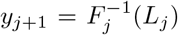. The infection times are ***y***^*′*^ = [*y*_1_, *y*_2_, *y*_3_, …, *y*_*j*_].

#### 3.4.1. Recovery of the information contained in y and T

To recover the evolution of the number of individuals in a particular compartment, we need to recover the times at which each individual leaves and enters that compartment. To achieve this, we concatenate [**y T**] and let **Q** the cumulative sum over rows. To follow the progression of individuals in some stage in the *k*-th column, *k >* 1, consider that the (*k −* 1)-th and *k*-th columns of **Q** contain the times when each individual entered and left that compartment respectively.

## 4. Examples

We can classify epidemic compartmental models in two categories: models in which no stage can be visited more than once, and models in which one or more stages can be visited more than once. The difference is that in the first case, there is a maximum for the number of transitions that an individual can make, thus, the size of a matrix containing the duration in each stage can be preset to *N × K*. In the second case the matrix can be of any size, presenting computational challenges. Here, we will deal only with the fist case and indicate how to proceed in the second case.

### 4.1. Example 1: and SEIQR model

The model in Figure 6 is a Susceptible-Latent-Infectious-Quarantined-Removed (*SEIQR*) model. Infectious individuals have contacts according a Poisson process with parameter *λ* and encounter susceptibles with probability *S/N*, thus, individuals leave the *S* compartment at a rate *λIS/N*. Once they leave the *S* compartment they go through the rest of stages *E, I, Q* and finally arrive to *R*, where they remain. The notation on top of each stage indicates the associated statistical distribution. For this model, the infectious contact rates are: ***λ***^*′*^ = [0, 2.1, 0] for stages *E, I* and *Q* respectively.

**Figure 6:**
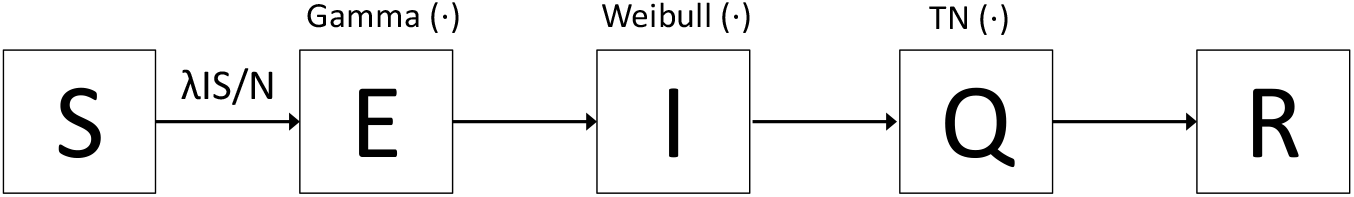
A Susceptible- Latent- Infectious- Quarantined-Removed (*SEIQR*) model. The notation on top of each stage indicates the associated statistical distribution.

In this case we use a Gamma(2.3, 2) distribution for the *E* stage, a Weibull (1, 2) for the *I* stage and a Gamma (5.7, 2.5) for the *Q* stage. Figure 7 shows a single simulation of the *SEIQR* for *N* = 10, 000. The Python code for this example (version 3.8.1) is provided as Supplementary material.

**Figure 7:**
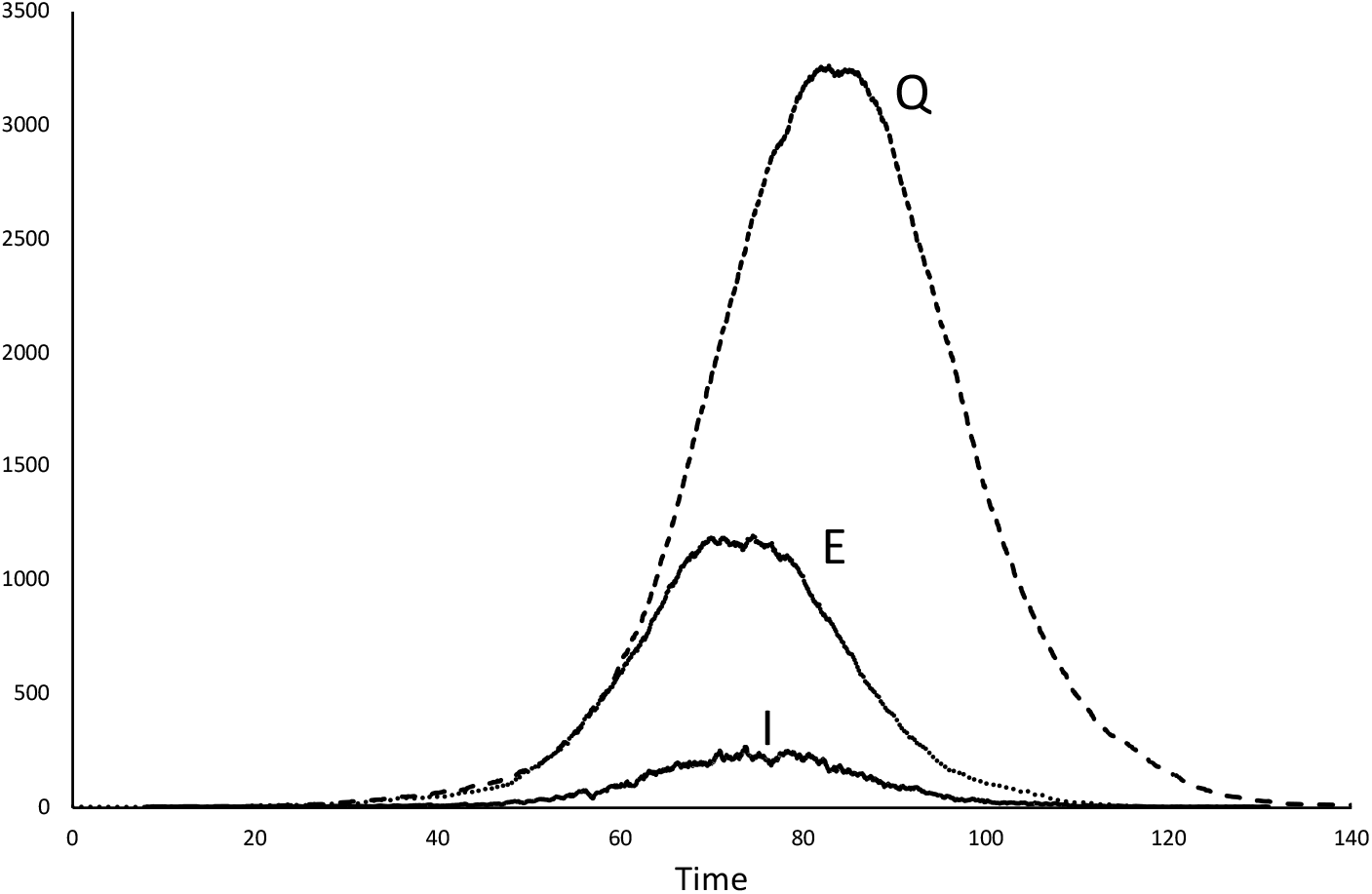
A simulation of the *SEIQR* model of Figure 6.

### 4.2. Example 2: a more general model

The model depicted in Figure 8 is an example where individuals are infectious at different stages and with different degree of infectiousness. This is an Ebola model where infectious individuals are infectious even while hospitalized or even dead before being buried, (Legrand et al., 2007). This is a complete example for the models in this category since not all stages may be visited and there are several kinds of infectious stages with differential degree of infectiousness. Here, the model has been modified to admit any distribution for the duration in stage *i, D*_*i*_(*•*).

**Figure 8:**
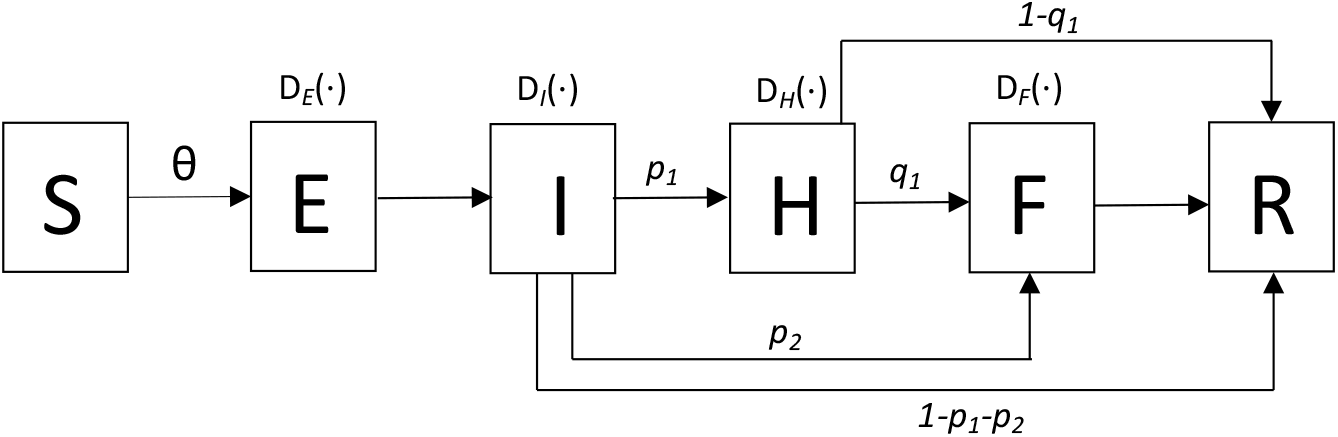
An Ebola transmission model adapted from Legrand et al. (2007), in which hospitalized and unburied are a source of infection. *S*, Susceptible individuals; *E*, Exposed individuals; *I*, Infectious; *H*, hospitalized; *F*, dead but not yet buried; *R*, removed. *Di*(*·*) indicates some general distribution for the duration in stage *i* described in Table 2. The total infection rate is Θ = (*λ*_*I*_ *I* + *λ*_*H*_ *H* + *λ*_*F*_ *F*)*S/N*.

**Figure 9:**
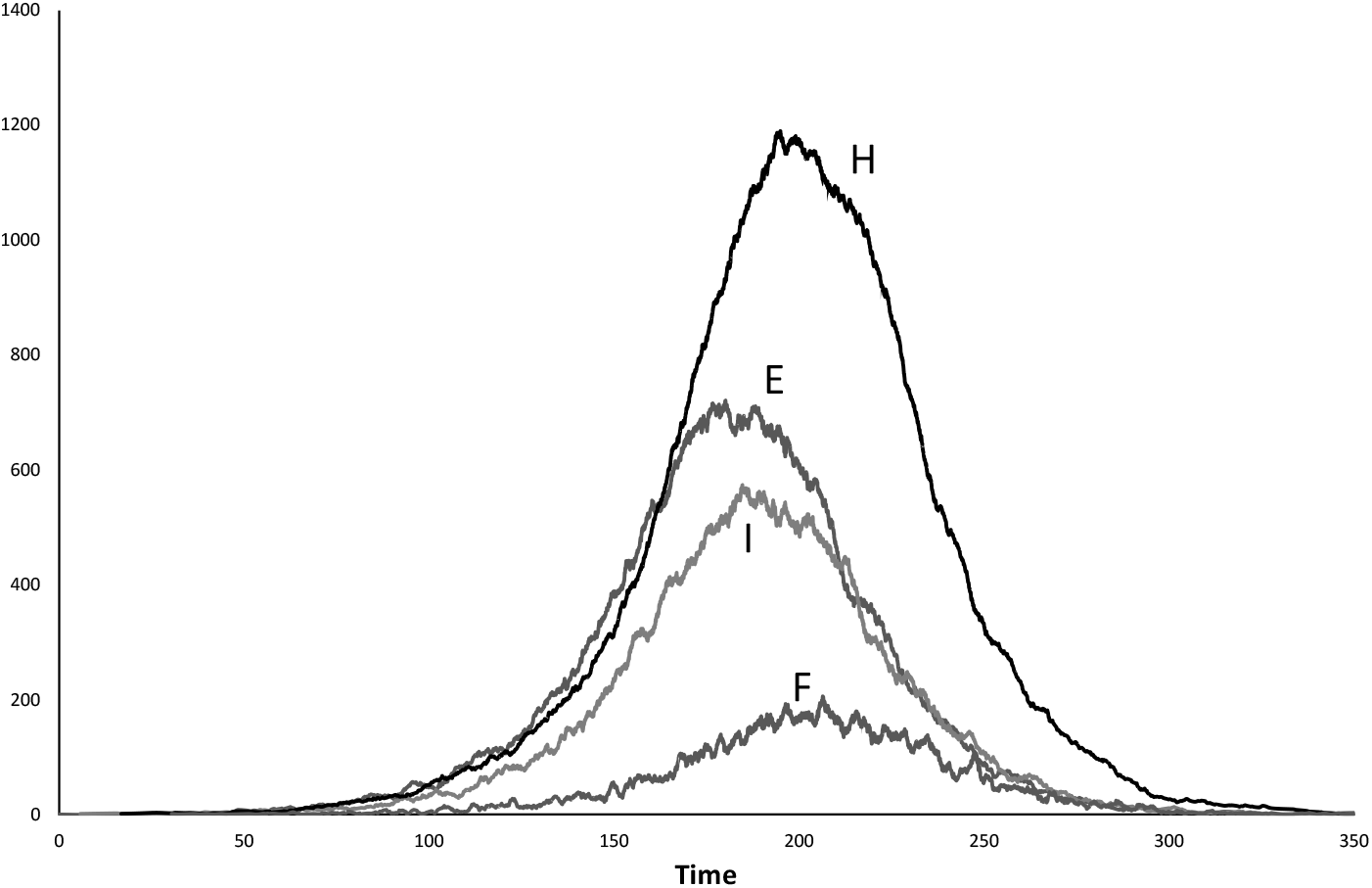
A single simulation of an Ebola epidemics depicted in Figure 8 in which hospitalized and unburied are a source of infection. The duration in each stage is described in Table 2.

We start by generating a matrix **T** with four columns, one for each of the stages *E, I, H* and *F*. Then, there are several strategies to simulate the possible paths that an individual can follow and here we chose a simple approach. Table 1 shows a list with the possible visited stages and the associated probabilities. We can simulate from this distribution and generate *N* possible paths which can then be recorded in a matrix **P** of 0’s and 1’s depending on the stages *{E, I, H, F }* being visited or not.

**Table 1:** Possible paths after infection with their probabilities.

| Path | Probability |
| --- | --- |
| $E - I - H - F$ | $p_1 q_1$ |
| $E - I - H$ | $p_1 (1 - q_1)$ |
| $E - I - F$ | $p_2$ |
| $E - I$ | $1 - p_1 - p_2$ |

**Table 2:** The distributions used to simulate the Ebola model from Legrand et al. (2007)

| Stage | Distribution | Parameters | Mean | SD |
| --- | --- | --- | --- | --- |
| $E$ | Truncated Normal in $[4,10]$ | $\mu = 7, \sigma = 3,$ | 7 | 1.62 |
| $I$ | Rayleigh | 4.3 | 5.39 | 2.81 |
| $H$ | Gamma | $\alpha = 7, \beta = 2.1$ | 14.7 | 5.55 |
| $F$ | Constant | 2 | 2 | 0 |

After the simulation of the *N* paths, the resulting matrix will look like:

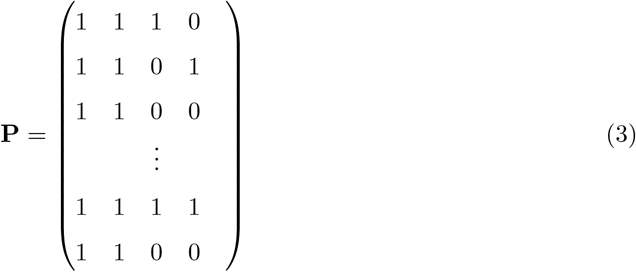

and then matrix **T** can be redefined as:

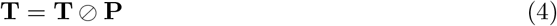

matrix **T** contains the time spent for each individual in each state, and we can apply the pseudocode suggested in *§*3.4 *to this matrix*.

#### 4.2.1. Example

We performed a single simulation of the Ebola model (Legrand et al., 2007) to illustrate this using arbitrary parameters and distributions for the duration on every state. For the model in Figure 8, we use *λ*_*I*_ = 0.15, *λ*_*H*_ = 0.1, *λ*_*F*_ = 0.05, *p*_1_ = 0.797, *p*_2_ = 0.163, *q*_1_ = 0.9, whereas we use the distributions indicated in Table 2 for each state. The infectious contact rates are: ***λ***^*′*^ = [0., 0.15, 0.1, 0.05] for stages *E, I, H* and *F* respectively and use *N* = 10, 000. The Python code is provided also as Supplementary material.

The first five rows of **T** are:

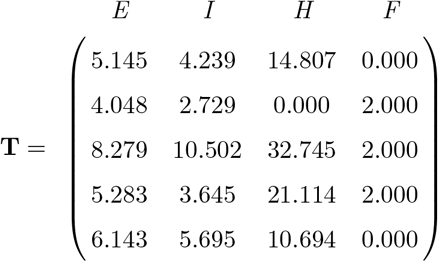

where we can see, for instance, that the first and fifth individuals were buried immediately, and the second died without being hospitalized.

## 5. Discussion

The purpose of this research is to introduce a method to simulate stochastic epidemics when sojourn times follow a general distribution. Using the true distribution may be useful to obtain better estimates of functionals of path integrals, for instance, to estimate the total bed-days required in hospitalization or in IC units. However, although it is clearly better than using a general exponential distribution or even an approximation to the first moments, it is important to note that we do not explore how much improvement is achieved because that would require to establish some optimality criteria that is beyond the scope of this work.

Models that allow one or more stages to be visited more than once can be simulated using the same theory, although they are clearly more complicated to implement in a computer because some rows of matrix **T** are longer than others. In practice, the major limitation of this method is computing time, which increases as a power of *N* : it takes on average of 0.7 seconds to generate a single simulation of the Ebola model in a 2.7 GHz MacBook Pro when *N* = 10^3^; 7 seconds in a population of *N* = 10^4^ and 1600 seconds when *N* = 10^5^. We found that about 92% of the total simulation time is required by the addition of the piecewise linear functions *F*_*j*_(*t*) = *F*_*j−*1_(*t*) + *f*_*j*_(*y*_*j*_, *t*), which is an area of opportunity.

Observe that models in which individuals move between compartments *i* and *j* according to some fixed probability *p*_*ij*_ implicitly consider that this probability is constant and independent of the individuals. It is reasonable to assume that *p*_*ij*_ was calculated using a frequentist approach, as it is customary in Markov models wit *p*_*ij*_ = *n*_*ij*_*/n*_*i*_ where *n*_*i*_ is the number of individuals that visited stage *i* and *n*_*ij*_ is the number of transitions between from stage *i* to *j*. If it happens that this probability depends on the length of stay in stage *i* the model is not Markovian anymore. An important advantage of the construction presented here is that it is possible to include a relaxation in the transition probabilities between compartments. For instance, in the Ebola model in Figure 8, suppose that the probability that an infected individual moves from stage *I* to stage *F* decreases the longer the individual stays in the *I* compartment, for example, following the relationship *e*^*−αt*^ where *t* is the time spent in stage *I*, then, we can construct vector **p**_1_ as a function of the residence times in stage *I*, **t**_*I*_ :

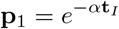

thus, building the matrix **P** in (3) with these considerations. Clearly, each row of **P** must be generated with information on the particular values of *p*_1_, *p*_2_ and *q*_1_ for each individual. The possibility of making individual transition probabilities opens the door to a new kind of models.

## Data Availability

No data available

## 7. Appendix 1

Let *X* be the first occurrence of a non-homogeneous Poisson process with parameter *λ*(*t*) following the piecewise set of linear functions:

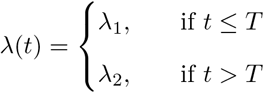

Observe that with probability 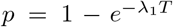, the event will occur in [0, *T*], and with probability 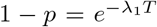 the event will occur after *T*. Thus *X* is a mixture distribution of two exponential distributions with parameters (*λ*_1_, *λ*_2_) and mixing parameters (*p*, 1 *− p*). One way to simulate this is by generating a random number *u* from a Uniform distribution and thus, *X* follows the same distribution than the mixture of *Y*_1_ = *−* log(*u*)*/λ*_1_ and *Y*_2_ = *−* log(*u*)*/λ*_2_. Now let *λ*_2_ = *kλ*_1_, thus, we have that *X* is the mixture distribution of:

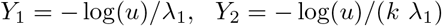

which reveals that we can simulate *X* by simulating an Exponential random variable *W* with parameter *λ*_1_, and if this is smaller than *T* we take *X* = *W* and if it is larger than *T*, we divide the excess *W − T* by *k* and add *T*. But this is equivalent to the construction of a function *F* (*t*) with the accumulated exposure provided by all infections by time *t* and and letting *X* = *F* ^*−*1^(*W*), where *W* is as described before. In our example:

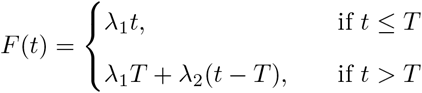

We can generalize this to a Poisson process with *λ*(*t*) defined as a piecewise set of linear functions in *r* intervals.

